# Critical Thinking and Ethical Decision-Making among Nursing Interns

**DOI:** 10.64898/2025.12.30.25343204

**Authors:** Wei Wang, Huina Xu, Yankai Shi

## Abstract

**Background:** Ethical decision-making is a core competency for nursing interns, supported by critical thinking as a central cognitive skill. However, the mechanisms linking these constructs remain unclear. This study examined the influence of critical thinking on ethical decision-making and the chain mediating roles of ethical sensitivity and sense of power.

**Methods:** A cross-sectional study was conducted from December 2024 to June 2025 among 429 nursing interns at a tertiary hospital in Zhejiang Province, using validated scales and structural equation modeling.

**Results:** Critical thinking, ethical sensitivity, sense of power, and ethical decision-making were positively correlated (P < 0.01). Ethical sensitivity and sense of power each mediated, and jointly serially mediated, the relationship between critical thinking and ethical decision-making.

**Conclusion:** Critical thinking enhances nursing interns’ ethical decision-making through ethical sensitivity and sense of power. Strengthening these factors in education may better translate cognition into ethical action.

## 1. Introduction

With advances in medical technology and an aging population, clinical practice increasingly presents challenges for nurses, including cognitive differences with patients, value conflicts, growing professional autonomy, and persistent imbalances between medical resource supply and demand. These factors contribute to frequent ethical decision-making (EDM) challenges in daily nursing practice (Faraco et al., 2022; Gonzalez-Garcia et al., 2025). The ability to apply nursing ethics effectively in complex situations and make patient-centered decisions has become an indispensable competency for nurses (Pritchard & Vieira-Moreno, 2025). EDM refers to the process by which nurses, guided by ethical theories and clinical experience, exercise professional judgment to select optimal actions (Barlow et al., 2018). Nurses’ EDM abilities not only influence the quality of care and professional ethical behavior (Alzghoul & Jones-Bonofiglio, 2020) but are also associated with professional values and coping with psychological stress (Chen et al., 2021; Yun et al., 2024). High-level EDM enables accurate identification of patients’ needs, reduces disputes, and enhances professional identity (Afenigus & Sinshaw, 2025). Nursing students, as the future clinical workforce, transition from theoretical learning to practice during internships, gradually shaping ethical awareness and reasoning. Surveys indicate that Chinese nursing interns generally have moderate EDM competence, and qualitative studies suggest insufficient ethical preparedness, potentially leading to breaches in confidentiality, neglect of privacy, and delayed or suboptimal decisions (Xu et al., 2025; Heidari et al., 2025). Identifying key factors influencing EDM is therefore essential for optimizing ethics education.

Among influencing factors, critical thinking (CT) is widely recognized as a core cognitive skill underlying clinical judgment and EDM. CT enables interns to analyze problems, compare solutions, and formulate rational, evidence-based judgments (Park et al., 2023). Empirical studies demonstrate a positive association between CT and EDM (Dang, Li, & Li, 2024; Silva et al., 2023). However, the mechanisms linking CT to EDM, particularly potential mediating pathways, remain underexplored. Ethical sensitivity (MS), the ability to recognize and interpret moral elements in situations, enhances EDM competence by allowing interns to identify dilemmas from the patient’s perspective and propose feasible strategies (Chen et al., 2021; Spekkink & Jacobs, 2021). Sense of power (SP), reflecting perceived influence and agency, affects emotional regulation and ethical behavior, with higher SP promoting confidence, adherence to principles, and accountability (He et al., 2025; Hwu & Pai, 2025). Evidence suggests higher MS can increase SP, indicating a potential serial mediation effect between CT and EDM (Fackler et al., 2015).

This study is grounded in Rest’s four-component model, which frames EDM as a multi-stage process from cognition to action (Spielthenner, 2008), and Trevino’s person–situation interaction model, which emphasizes the joint influence of individual traits and situational factors (Trevino, 1986). Within this framework, CT provides the cognitive foundation, MS enables recognition of ethical conflicts, and SP facilitates translating ethical intention into action. Accordingly, this study investigates the cognitive–affective–behavioral chain underlying nursing interns’ EDM to inform nursing ethics education and professional development, as shown in Figure 1:

**Figure1.**
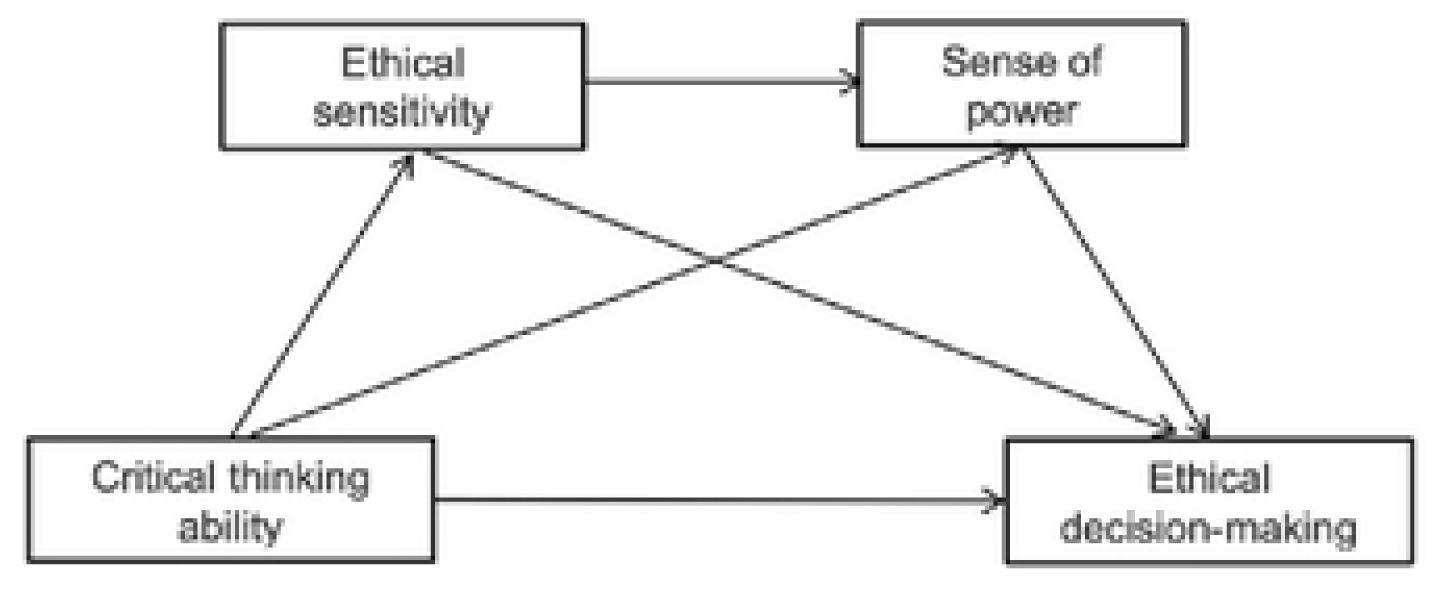
Hypothetical structure model.

H1: Nursing interns’ critical thinking ability (CT) positively predicts their ethical decision-making competence (EDM).

H2: Ethical sensitivity mediates the relationship between CT and EDM.

H3: Sense of power mediates the relationship between CT and EDM.

H4: Ethical sensitivity and sense of power jointly mediate the relationship between CT and EDM through a serial pathway.

## 2. Methods

### 2.1 Study Design

This study employed a cross-sectional design and was conducted at a tertiary hospital in Ningbo, Zhejiang Province, from December 2024 to June 2025. We recruited nursing students undertaking clinical internships using a convenience sampling approach. Following Kendall’s principle for sample size estimation, we based the calculation on the Critical Thinking Ability Scale, which had the largest number of items among all instruments. A sample size of 5–10 times the number of items was recommended(Ni, 2010), yielding a minimum of 350 participants. Considering a 20% potential attrition rate, we aimed to collect at least 420 questionnaires. A total of 450 questionnaires were distributed, of which 429 were completed validly, yielding an effective response rate of 95.3%. The study complied with the Declaration of Helsinki and obtained approval from the Ethics Committee of the participating hospital (Approval No.: Li Huili Hospital Ethics Review No. 407 (2025 Research)). All participants provided informed consent and participated voluntarily. We included students who were currently enrolled in the nursing program, undergoing clinical internships at the tertiary hospital, and voluntarily agreed to participate with signed informed consent. We excluded students with less than one month of internship experience or those who submitted incomplete questionnaires or provided responses with obvious logical inconsistencies.

### 2.2 Data Collection

We collected data through an online questionnaire hosted on the Wenjuanxing platform (https://www.wjx.cn/). The research team presented the study objectives, content, and instructions on the questionnaire homepage, emphasizing voluntary participation, anonymity, and strict data confidentiality. Under the guidance of clinical instructors, participants accessed the questionnaire via QR code and completed it online. To ensure completeness, the system allowed only one submission per IP address and required participants to answer all items before submission. The questionnaire included demographic characteristics, critical thinking ability, ethical sensitivity, sense of power, and ethical decision-making.

### 2.3 Quality Control

To ensure measurement validity, we selected instruments that have been widely validated both domestically and internationally. After data collection, the research team reviewed all questionnaires and excluded responses exhibiting:(1) Consistent response patterns, such as selecting the same option for all items except demographics; (2) Logical inconsistencies or clearly implausible answers; (3) Completion times under two minutes, indicating insufficient attention.

### 2.4 Measures

#### 2.4.1 Demographic Questionnaire

We designed a self-developed demographic questionnaire based on previous studies, which included seven items such as gender, academic year, interest in nursing as a major, and whether the participant had attended a nursing ethics course. The instrument served to collect basic demographic and background information of the participants.

#### 2.4.2 Judgments about Nursing Decisions (JAND)

We assessed ethical decision-making ability using the Judgments about Nursing Decisions (JAND) scale developed by Ketefian(Ketefian, 1981). Zhu introduced and revised the scale for use in China in 2011 to better align with local nursing ethics practice(Zhu, 2011). The instrument consists of six cases and two situational questions, covering two dimensions: ethical cognition and ethical behavior, with a total of 48 items. The overall Cronbach’s α coefficient was 0.974, and values for the two dimensions ranged from 0.949 to 0.959, indicating excellent reliability and validity.

#### 2.4.3 Personal Sense of Power Scale

The Personal Sense of Power Scale by Anderson et al. (Anderson et al., 2012)was employed to evaluate participants’ perceived power. The instrument contains eight items—four reflecting high power and four reflecting low power—using a seven-point Likert scale, where higher scores represent greater perceived power. Reverse scoring was applied to items 2, 4, 6, and 7. The scale showed strong reliability in this study (Cronbach’s α = 0.911).

#### 2.4.4 Ethical Sensitivity Questionnaire for Nursing Students (ESQ-NS)

To assess ethical sensitivity, we utilized the ESQ-NS developed by Muramatsu et al.(Muramatsu et al., 2019). Liu et al.(Liu, 2022) translated and revised the instrument into Chinese in 2022. The scale consists of 13 items across three dimensions, rated on a four-point scale. Higher scores indicate stronger ethical sensitivity. All items were designed around ethical issues commonly encountered by nursing students in clinical practice, making the scale particularly suitable for this population. Reliability analysis indicated that the Chinese version had a Cronbach’s α of 0.886, reflecting satisfactory internal consistency.

#### 2.4.5 Critical Thinking Disposition Inventory–Chinese Version (CTDI-CV)

Critical thinking ability was assessed using the CTDI-CV, originally developed by Facione et al. and subsequently translated and revised by Peng et al.(Peng, 2004) The inventory comprises 70 items across seven dimensions, with 10 items per dimension, rated on a six-point Likert scale, where higher scores reflect stronger critical thinking disposition. A total score ≤210 suggests a weak disposition, 211–279 indicates a moderate disposition, ≥280 reflects a strong disposition, and ≥350 indicates a markedly positive disposition. The Chinese version demonstrated excellent internal consistency, with a Cronbach’s α of 0.984.

### 2.5 Statistical Analysis

We performed all statistical analyses using IBM SPSS Statistics 25.0 and AMOS 29.0, while Excel was used to organize and manage the raw data. (1) Reliability and Validity Testing: We calculated Cronbach’s α coefficients for each scale to assess reliability. CFA was conducted in AMOS 29.0 to examine construct validity. We evaluated model fit using the CFI, NFI, TLI, GFI, RMSEA, SRMR, and χ^2^/df, with acceptable thresholds defined as RMSEA < 0.08, SRMR < 0.08, CFI, TLI,NFI, GFI > 0.90, and χ^2^/df < 5. (2) Common method variance was examined through Harman’s single-factor test and a single-factor confirmatory factor analysis in AMOS. These procedures ensured the objectivity and reliability of the data. (3) Descriptive and Inferential Statistics: We used descriptive statistics to summarize participants’ demographic characteristics and the distribution of main study variables. The assessment of demographic variations in ethical decision-making was performed using independent-sample t-tests and one-way ANOVA.

We further applied Spearman correlation analysis to explore associations among critical thinking, ethical sensitivity, sense of power, and ethical decision-making. (4) Mediation Analysis: to test the hypothesized serial mediation model, we constructed a structural equation model in AMOS 29.0. We assessed the mediation effects of ethical sensitivity and sense of power on the association between critical thinking and ethical decision-making using 5,000 bootstrap resamples. A 95% CI that excluded zero indicated a significant mediation effect.

## 3. Results

### 3.1 Demographic Characteristics

A total of 429 nursing interns were included in this study, comprising 333 females (78.0%) and 96 males (22.0%). The sample consisted of 203 third-year students (47.3%) and 226 fourth-year students (52.7%). Regarding family structure, 58 participants (13.5%) were only children, while 371 (86.5%) were not. In terms of residence, 253 students (59.0%) came from urban areas, and 176 (41.0%) from rural areas. Concerning professional identity, 149 participants (34.7%) reported liking nursing, 202 (47.1%) expressed neutrality, and 78 (18.2%) reported disliking it. With respect to nursing ethics education, 253 students (59.0%) had received ethics-related courses, while 176 (41.0%) had not. The primary sources of ethics knowledge included the internet (n = 225, 52.4%), school (n = 158, 36.8%), and family (n = 46, 10.7%). The specific information is shown in Table 1.

### 3.2 Descriptive and Correlational Analyses

The mean scores of nursing interns were as follows: ethical decision-making 238.31±33.14,critical thinking 238.44±39.29,ethical sensitivity 38.12± 5.37, and sense of power 31.64 ± 6.03. Spearman correlation analysis revealed that critical thinking was positively associated with ethical decision-making and its two subdimensions (r =0.363–0.390, *P*<0.001). Ethical sensitivity and its dimensions were also positively correlated with Ethical decision-making and its dimensions (r=0.131–0.492, *P* < 0.001). Furthermore, sense of power showed significant positive correlations with ethical decision-making and the ethical behavior dimension (r=0.105–0.166, *P*< 0.01). A positive correlation was observed between ethical sensitivity and sense of power, with the strongest associations found in the “respect for individuals” dimension (r =0.191–0.396, *P*< 0.01). See Table 2 for details.

### 3.3 Mediating effect tests

#### 3.3.1 Common Method Bias

The assessment of common method bias was carried out using Harman’s single-factor test. The analysis yielded 26 factors with eigenvalues above 1, and the first unrotated factor explained 34.83% of the variance, which falls below the 40% cutoff, indicating minimal concern regarding common method bias.

#### 3.3.2 Model Fit of the Structural Equation Model

We constructed a serial mediation model in AMOS 26.0, with critical thinking as the independent variable, ethical sensitivity as mediator 1, sense of power as mediator 2, and ethical decision-making as the dependent variable. Mediation effects were examined using bootstrap resampling with 5,000 iterations and 95% confidence intervals. The modified model showed good fit: χ²/df = 3.681 (<5), RMSEA = 0.079 (< 0.08), and CFI, TLI, and IFI all exceeded 0.90, suggesting satisfactory model fit. The model is shown in Figure 2.

**Figure 2.**
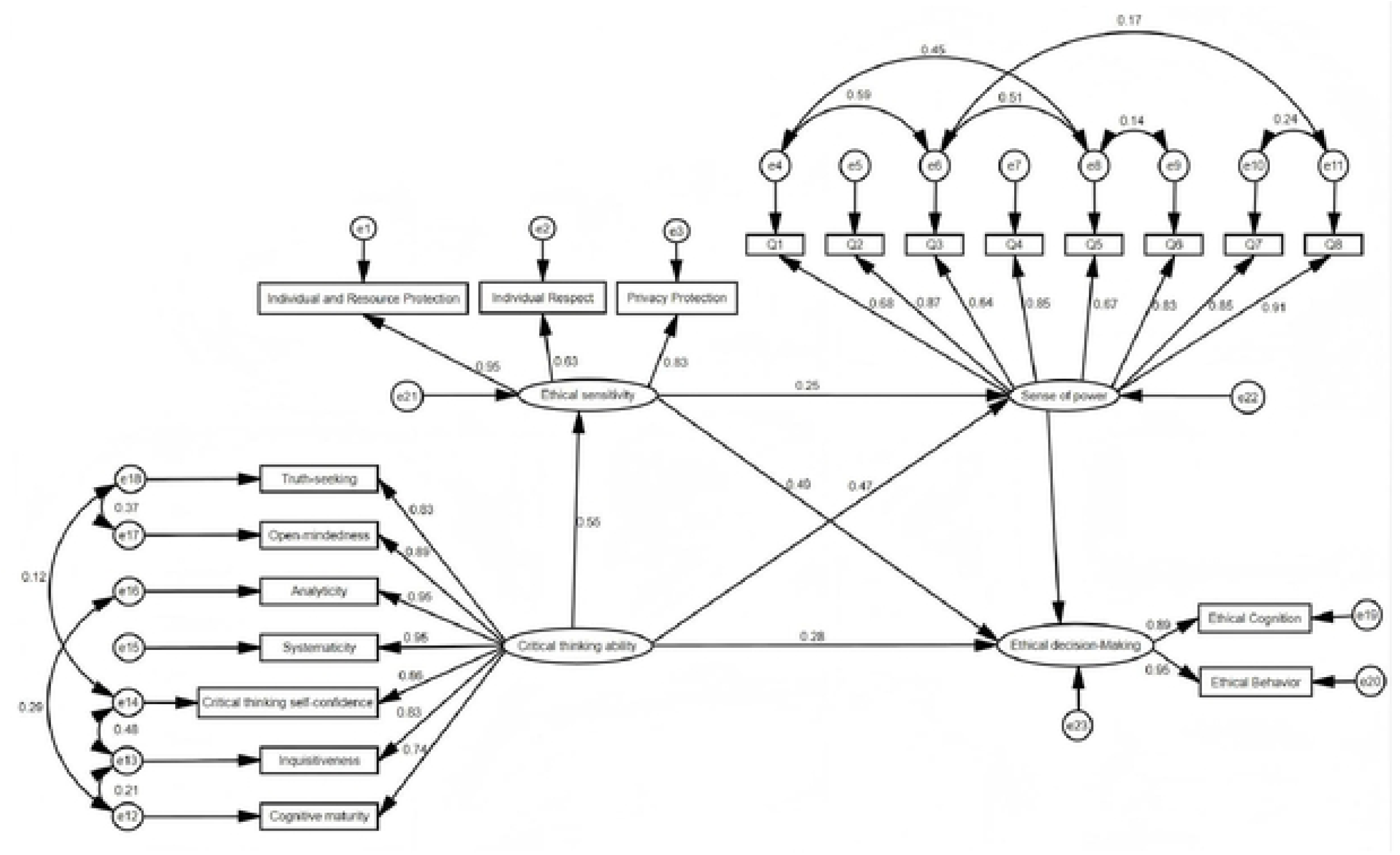
The chain mediation model of critical thinking, ethical sensitivity, sense of power, and ethical decision-making.

#### 3.3.3 Path Coefficients

The analysis revealed that all path coefficients were significant, with p-values below 0.05, as shown in Figure 2. Critical thinking had a direct effect on ethical decision-making (β = 0.282) and also positively predicted ethical sensitivity (β = 0.547) and sense of power (β = 0.465). Ethical sensitivity positively predicted sense of power (β = 0.246). Both ethical sensitivity (β = 0.496) and sense of power (β = 0.251) significantly predicted ethical decision-making. These outcomes indicate that critical thinking impacts ethical decision-making both directly and through the chain mediating roles of ethical sensitivity and sense of power.

#### 3.3.4 Chain mediating effect

Three indirect pathways were significant. The strongest effect was observed in the pathway “critical thinking → ethical sensitivity → ethical decision-making” (effect size = 0.876), accounting for 38.4% of the total effect, highlighting ethical sensitivity as a core mediator. The pathway “critical thinking → sense of power → ethical decision-making” yielded an effect size of 0.382 (16.8% of the total effect), indicating that sense of power also played a mediating role. The serial pathway “critical thinking → ethical sensitivity → sense of power → ethical decision-making” had an effect size of 0.112, representing 4.9% of the total effect. Although smaller in magnitude, this effect remained statistically significant, as shown in Table 3.

## 4. Discussion

### 4.1 Mediating Role of ethical sensitivity

This study found that nursing interns demonstrated moderately high levels of critical thinking and ethical sensitivity, slightly higher than those reported in some previous studies(Nemati-Vakilabad et al., 2023; Wen et al., 2025). This difference may be attributed to sample characteristics, as most participants were female and enrolled in their third or fourth year of study. Prior research(Alnajjar Ph & Abou Hashish Ph, 2021; Dang, Li, Li, et al., 2024) has shown that both gender and academic year influence ethical awareness and critical thinking, with female and senior students often displaying greater sensitivity in ethical recognition and judgment. As clinical experience accumulates, students become more adept at identifying and analyzing complex problems in real care settings, thereby gradually improving their critical thinking and ethical sensitivity. More importantly, Our findings confirmed that ethical sensitivity mediates the relationship between critical thinking and ethical decision-making ability, which is consistent with findings reported by Chen et al.(Chen et al., 2021), who demonstrated that the level of ethical sensitivity determines whether individuals can promptly recognize and respond to ethical dilemmas in clinical practice, facilitating the translation of critical thinking into higher-order ethical judgment and decision-making. Nursing students with stronger critical thinking skills are more likely to detect potential ethical issues in complex situations, which enhances their ethical sensitivity and, in turn, strengthens their ethical decision-making. As the cognitive starting point of ethical decision-making, ethical sensitivity serves as a key psychological mechanism enabling students to make scientific and rational decisions when faced with value conflicts. Alnajjar et al.(Alnajjar Ph & Abou Hashish Ph, 2021; Yi et al., 2024) also reported that nursing interns generally demonstrate moderately high levels of ethical sensitivity, suggesting that this ability is established during academic training but still requires reinforcement through curricular interventions and clinical experiences. Therefore, nursing education should prioritize strategies such as scenario simulation, case-based discussions, and reflective learning to help students continuously recognize and differentiate ethical dilemmas in authentic or simulated contexts, thereby enhancing ethical sensitivity and strengthening ethical competence.

### 4.2 Mediating Role of Sense of Power

An individual’s sense of power represents their perceived capacity for influence and agency in social interactions or particular situations, which directly affects their responsibility, emotional regulation, and confidence in ethical decision-making(Hwu & Pai, 2025). This study showed that nursing interns reported a moderate level of sense of power, suggesting that while they experience some autonomy in clinical practice, they may also face risks of insufficient self-efficacy. Previous research(Ahn & Choi, 2015) has emphasized that an adequate sense of power enables nursing students to remain calm and adhere to ethical principles in high-pressure contexts, whereas low levels may weaken their agency and ethical performance. Our findings further validated the mediating role of sense of power between critical thinking and ethical decision-making ability. Kadosh and colleagues (Kadosh & Rozani, 2025)highlighted the positive influence of clinical empowerment, noting that students with stronger critical thinking are more likely to develop autonomy and agency in practice, which enhances their sense of power. This enhanced sense of power, in turn, enables them to remain confident and composed when facing ethical conflicts, thereby making rational and ethically sound decisions. These results are also consistent with He et al.(He et al., 2025), who argued that a strong sense of power promotes nurses’ ethical efficacy and decision implementation. Accordingly, nursing educators and clinical mentors should adopt strategies such as constructive feedback, role empowerment, and collaborative learning to strengthen students’ sense of power. Such approaches not only reinforce students’ professional identity and confidence but also facilitate the transformation of critical thinking into effective ethical decision-making.

### 4.3 Chain Mediating Effect of ethical sensitivity and Sense of Power

Ethical decision-making, according to Rest’s four-component framework(Spielthenner, 2008), proceeds through stages of ethical sensitivity, moral judgment, moral character and moral motivation. Trevino’s person–situation interactionist perspective further underscores the dynamic interplay between personal characteristics and contextual factors in guiding ethical behavior(Trevino, 1986). In this study, critical thinking emerged as a core cognitive trait that enables analytical reasoning, while sense of power functioned as a contextual perception that influences responsibility, control, and self-efficacy in ethical challenges. The results demonstrated that critical thinking not only exerted direct effects on ethical decision-making but also influenced it indirectly through chain pathways. This indicates that ethical sensitivity and sense of power do not operate independently but instead form a continuous mechanism. Specifically, higher levels of critical thinking help students more quickly recognize ethical issues in clinical settings, thereby enhancing ethical sensitivity. Heightened ethical sensitivity then reinforces their sense of responsibility and perceived influence in ethical contexts, which strengthens their sense of power, ultimately promoting ethical decision-making. Although the effect size of this chain mediating effect was relatively small, it remained statistically significant. These findings extend beyond prior studies that only considered single mediators(Turan & Çekiç, 2023), offering a more comprehensive framework for understanding the psychological mechanisms underlying ethical decision-making among nursing students.

Nevertheless, this study also found that overall ethical decision-making ability among nursing interns was at a moderate-to-low level, lower than the results reported by Sari et al. (Sari et al., 2018)and Xu et al.(Xu et al., 2025) Notably, scores for ethical cognition were higher than those for ethical behavior, suggesting that while students can identify ethical problems at the theoretical level, they often face difficulties in translating cognition into action. This gap between ethical cognition and behavior has also been observed in other studies(Turan & Çekiç, 2023), and may be explained by role limitations in clinical settings, lack of practical experience, and concerns about potential ethical consequences. Therefore, nursing education must not only cultivate critical thinking and ethical sensitivity in classroom instruction but also bridge the cognition–behavior gap through clinical simulations, guided mentoring, and group discussions. Furthermore, creating empowering learning environments in clinical teaching can help students strengthen both responsibility and sense of power, thereby enabling them to transform critical thinking into ethical action and ultimately enhance their ethical decision-making competence.

## 5. Conclusion

This study confirmed that critical thinking significantly influences nursing interns’ ethical decision-making ability, both directly and indirectly through the chain mediating effect of ethical sensitivity and sense of power. The findings highlight ethical sensitivity as a crucial factor in recognizing and interpreting ethical dilemmas, while sense of power plays a key role in transforming ethical cognition into concrete decisions and actions. These findings highlight the necessity of incorporating both cognitive and affective components into education and clinical practice. The contribution of this study lies not only in elucidating the internal mechanisms of ethical decision-making among nursing students but also in offering theoretical and practical insights for nursing ethics education. Future efforts should focus on developing more targeted teaching and support strategies to cultivate highly qualified nurses with strong critical thinking, ethical sensitivity, and a sense of responsibility, thereby enabling them to effectively cope with complex and dynamic ethical challenges in clinical practice.

### Limitations

First, the study sample was primarily drawn from nursing interns at one hospital, which may constrain the generalized ability of the results. Expanding future research to encompass students from different regions, institution types, and cultural contexts could enhance the applicability of the findings. Second, data were mainly collected using self-report questionnaires. Although the instruments employed demonstrated good reliability and validity, reliance on self-report measures may compromise the objectivity of the results. To enhance the robustness and credibility of future findings, incorporating multi-source data—such as mentor evaluations, performance in simulated scenarios, or peer feedback—is recommended. Finally, this study focused on the mediating roles of ethical sensitivity and sense of power. Nevertheless, ethical decision-making in nursing is a multifaceted process encompassing cognitive, emotional, and behavioral components, and it may also be shaped by contextual factors such as ethical climate and organizational culture. The integration of these variables into future models would allow for the development of a more comprehensive theoretical framework.

## Data Availability

All relevant data are within the manuscript and its Supporting Information files.

## Acknowledgments

The contributions of the nursing students who participated in this research are sincerely appreciated. Their enthusiastic involvement and honest feedback were invaluable for data collection and contributed to the reliability of the results.

## Author Contributions

Wei Wang and Huina Xu conceived and designed the study. We Wang performed data analysis and drafted the manuscript. Huina Xu contributed to data collection and assisted with manuscript revision. Yankai Shi supervised the study, provided critical revisions, and approved the final version of the manuscript. All authors read and approved the final manuscript.

## Data Availability

Access to the datasets generated and analyzed during this study is available from the corresponding author upon reasonable request.

## Ethical Approval and Consent to Participate

This study was conducted in accordance with the principles of the Declaration of Helsinki and received ethical approval for the study protocol from the Medical Ethics Committee of Ningbo Medical Centre Lihuili Hospital (Approval No. Li Huili Hospital Lunshen 2025 Research No. 407). All participants provided written informed consent before their enrollment in the research.

